# Unilateral and Bilateral Theta Burst Stimulation for Treatment-Resistant Depression: Follow up on a Naturalistic Observation Study

**DOI:** 10.1101/2024.05.19.24307592

**Authors:** Mariam Elnazali, Ashan Veerakumar, Mervin Blair, Emily L. Pearce, Noah Kim, Sreya Sebastian, Jonathan B. Santo, Iouri E. Rybak, Amer M. Burhan

## Abstract

**Objectives:** Theta burst stimulation (TBS) is a novel and faster modality of transcranial magnetic stimulation, which is showing promise as a treatment-resistant depression (TRD) treatment. Though TBS can be applied unilaterally or bilaterally, few studies have compared the effectiveness of both approaches in a naturalistic clinical sample. In this retrospective chart review, we aimed to: (1) replicate previous bilateral sequential TBS effectiveness in a larger cohort of patients at a single centre, (2) present treatment outcome data between unilateral and bilateral TBS approaches, (3) investigate baseline factors associated with our observed outcomes, and (4) examine the sustainability of response, with follow-up data up to 6 months from patients.

**Methods:** We included 161 patients who received TBS (unilateral: n = 64 (40%), 45.55 ± 14.25 years old, 55% females; bilateral: n = 97 (60%), 47.67 ± 15.11 years old, 58% females).

**Results:** Firstly, we observed 47% response and 34% remission in the bilateral group, replicating findings from a smaller naturalistic study from our group; patients receiving unilateral TBS displayed 36% response and 26% remission, with no significant differences found between unilateral and bilateral TBS in remission and response rates. Secondly, bilaterally stimulated patients needed fewer treatments than those stimulated unilaterally (27 vs 29 on average respectively, *t* [159] = 3.31, *p* = .001), and had significantly lower anxiety symptoms post treatment (GAD-7) relative to patients receiving unilateral stimulation, *F* (1,148) = 3.95, *p* =0.049. Thirdly, no baseline factors were found to predict treatment outcomes. Lastly, after six months, 69% of patients who met the response criteria did not require additional treatment or a change in medication.

**Conclusions:** Our findings support the efficacy and tolerability of TBS in TRD and indicate that bilateral TBS may have a superior anxiolytic effect and offer a slightly faster time to response.

## Introduction

Theta burst stimulation (TBS) is a patterned form of transcranial magnetic stimulation that involves delivering triplet bursts of gamma (i.e., 50 Hz) frequency pulses every 200 milliseconds (at theta frequency i.e., at 5 Hz). This pattern of stimulation has been shown to be efficient and capable of inducing a neuroplastic effect. Intermittent TBS (iTBS) involves the application of TBS for 2 seconds with an 8-second break and has shown to be excitatory and likely to induce long-term potentiation, while continuous TBS (cTBS) is largely inhibitory and likely induces long-term depression (LTD).^1^ In therapeutic applications for depression, iTBS has been mainly applied to the left dorsolateral prefrontal cortex; this 3-minute paradigm has been shown to be non-inferior to the 37-minute, high frequency repetitive transcranial magnetic stimulation (rTMS) approved by the FDA to treat treatment-resistant depression (TRD).^2^

Bilateral sequential TBS (bsTBS) involves applying cTBS to the right PFC (rcTBS) followed by iTBS to the left PFC (liTBS). A study by Li et al. (2014)^3^ compared different TBS protocols and found bsTBS to have possibly superior patient outcomes to liTBS, though no significant statistical differences were found. Furthermore, Mutz et al. (2019)^4^ conducted a network meta-analysis to compare the efficacy and acceptability of non-surgical brain stimulation treatments including electroconvulsive therapy (ECT) and rTMS, finding a higher odds ratio for bsTBS compared to liTBS (∼4 vs. ∼3). However, no significant difference was observed between the two treatments when directly compared.^4^ More recently bsTBS was studied in comparison to bilateral standard rTMS in older adults and was found to be non-inferior.^5^

We previously reported on the safety and efficacy of bsTBS in a clinic cohort of 50 patients with highly resistant depression. We found a 28% remission rate (Hamilton Depression Rating Scale (HAMD-17) score of ≤7) and a 52% response rate (≥50% reduction in HAMD-17).^6^ We also found this paradigm to be safe and efficacious in a sub-sample of patients with TRD co-morbid with military related posttraumatic stress disorder.^7^

Studies into the impact of individual factors on the efficacy of rTMS remain limited. Fitzgerald et al. (2016)^8^ conducted a study exploring response patterns of rTMS, examining response pattern, rate of response, remission, and potential clinical and demographic predictors of treatment response; their findings revealed that 46% of participants responded and 31% remitted, following treatment with rTMS. Having less severe depression scores at baseline and shorter and recurrent depression episodes (rather than single episodes) produced a greater likelihood of treatment response. In addition, a study conducted by Fregni et al. (2006)^9^ attempted to examine individual predictors of rTMS treatment efficacy, finding predictors that included younger age and being less treatment resistant.

In older adults, Valiengo et al. (2022)^10^ systematically reviewed and meta-analyzed evidence of rTMS efficacy in major depressive disorder (MDD). Contrary to past findings, there was a significant correlation with older age and a higher number of TBS treatments, with a more significant improvement in depression scores.^10^ In their multi-centre study, Bouaziz et al. (2023)^11^ investigated individual predictors of improved treatment outcomes, finding that a greater improvement in depressive symptoms was associated with more severe pretreatment scores.^11^

Despite promise from small studies^6^ and non-inferiority in older adults in a large study,^5^ further research is needed to evaluate the efficacy of TBS in treating MDD. Lastly, there is a lack of studies examining the long-term sustainability of treatment response.

In a naturalistic brain stimulation setting we aimed to: (a) examine the replicability of previous findings of efficacy for bsTBS in a larger cohort of patients, (b) directly compare treatment outcomes between unilateral and bilateral TBS approaches, (c) examine individual predictors of treatment efficacy; and (d) investigate sustainability of TBS, by examining follow-up data six months post treatment for patients that met response criteria.

## Methods

### Sample and outcome measures

This retrospective study has been approved by the Western University office of Human Research Ethics Board at St. Joseph’s Health Care London in compliance with the Declaration of Helsinki.

Data was collected retrospectively, following patient chart reviews, and as such, informed consent was not required for data collection. All patients had consented to treatment through the TBS clinic at Parkwood Institute Mental Health Care Facility between May 1, 2015, and June 15, 2020. The inclusionary criteria for the study were that patients needed to be between the ages of 18-105 years old, have a diagnosis of TRD, and their first course of treatment had to be their first course of adequate sessions (≥ 20 sessions). 161 patients between the age of 19-87 years of age were included in the study, completing a range of 20-30 TBS sessions. Of the 16 total patients who were excluded from the study, 15 did not complete an adequate first course of sessions, and one patient received standard rTMS treatment.

Pre- and post-psychiatric scores were collected from four different psychiatric scales: the 17-item HAMD-17,^12^ the Clinical Global Impression Scale (which includes both the CGI-S and the CGI-I),^13^ the 9-item Patient Health Questionnaire (PHQ-9),^14^ and the 7-item Generalized Anxiety Disorder scale (GAD-7)^15^.

#### 2.2. Data analysis

Data analyses were conducted using SPSS statistics version 27. To examine the four study objectives, first, bilateral response and remission rates were compared to those reported by Burhan et al. (2020)^6^ using chi-square analysis.

Second, we performed independent sample t-tests for differences in baseline treatment scores and demographics for the HAMD-17, GAD-7, PHQ-9, and CGI-S, as well as age and number of TBS sessions between unilateral and bilateral patients. A chi-square test of independence was performed to examine the relationship between gender and stimulation type. A comparison of post-treatment scores for the HAMD-17, PHQ-9, GAD-17, and CGI-I was examined between the two types of stimulation groups in the current study. Specifically, a one-way analysis of covariance (ANCOVA) was conducted to examine whether there were differences in post HAMD-17, GAD-7, and the PHQ-9 scores between unilateral and bilateral TBS, while controlling for baseline differences. Lastly, for scores on the CGI, a chi-square test of independence was conducted to examine whether there were differences between unilateral and bilateral TBS. Chi-square tests were also conducted between stimulation groups to examine if there were differences in response and remission rates. To examine differences in side effects between unilateral and bilateral stimulation, a multivariate analysis of variance (MANOVA) was conducted, which included muscle contractions, pain or discomfort, scalp irritation, syncope, heart rate drop, heart rate rise, post diastolic drop, post diastolic rise, post systolic drop, and post systolic rise.

Third, a structural equation modeling (SEM) analysis was conducted to investigate individual predictors of TBS efficacy. The SEM model included age, gender, number of sessions, and type of stimulation as predictors. The SEM allowed us to examine multiple outcomes at the same time for the HAMD-17, PHQ-9, GAD-7, and CGI, while also controlling for shared variances between predictors and multiple outcomes; it also enabled us to investigate any moderating influence of the psychiatric measures and covariates (age, gender, number of sessions and TBS side).

Lastly, we evaluated the sustainability of TBS by examining patients who responded to the treatment six months later. The data was analyzed to determine whether any additional treatment had been required, such as a second TBS treatment or a course of ECT or a change in medication.

### Treatment

A TMS Magstim Super Rapid 2 machine (The Magstim Company Ltd, TM., UK) was utilized in a brain stimulation clinic to successively administer rcTBS at the F4 location of the international 10-20-20 EEG localization system (right dlPFC), followed by liTBS at the F3 location (left dlPFC) for patients receiving bilateral TBS. Patients receiving unilateral TBS only received iTBS at the F3 location (left dlPFC). At each site, 600 pulses were delivered in bursts varying between 40-50 Hz at a rate of 5Hz (theta range) to allow treatment at 100% of the resting motor threshold (RMT) (average of 45%), as established by induction of a visible motor response in the hypothenar hand muscle in 3/5 trials. A nurse-administered questionnaire was used to report any adverse effects during and after each treatment, which were used to assess tolerability.

## Results

Of the patients included in this naturalistic clinical study, 161 received TBS (47.41 ± 13.89 years old; 57% female), with 64 (40%) receiving unilateral TBS (45.55 ± 14.25 years old, 55% female) and the remaining 97 (60%) receiving bilateral TBS (47.67 ± 15.11 years old, 58% female).

Following completion of their TBS treatment course (unilateral: mean 29 sessions ± 2.80; bilateral: mean 27 sessions ± 4.55) 49/157 patients (31.21%) achieved remission and 67/157 participants responded to treatment for a total response rate of 42.68%. Four patients had missing HAMD-17 scores and were excluded from the response and remission rate calculations.

Regarding the first objective, the bsTBS remission and response rates in our study (34.38% (33/96) and 46.88% (45/96) respectively) were comparable to those reported by Burhan et al (2020) (28% and 52%, respectively), with no significant differences found between these rates across the two studies (X^2^ (1, N= 146) < 1, *p* > .05). To address the second objective, we compared demographics and baseline scores between the unilateral and bilateral cohort (see Table 1) .

**Table 1.**
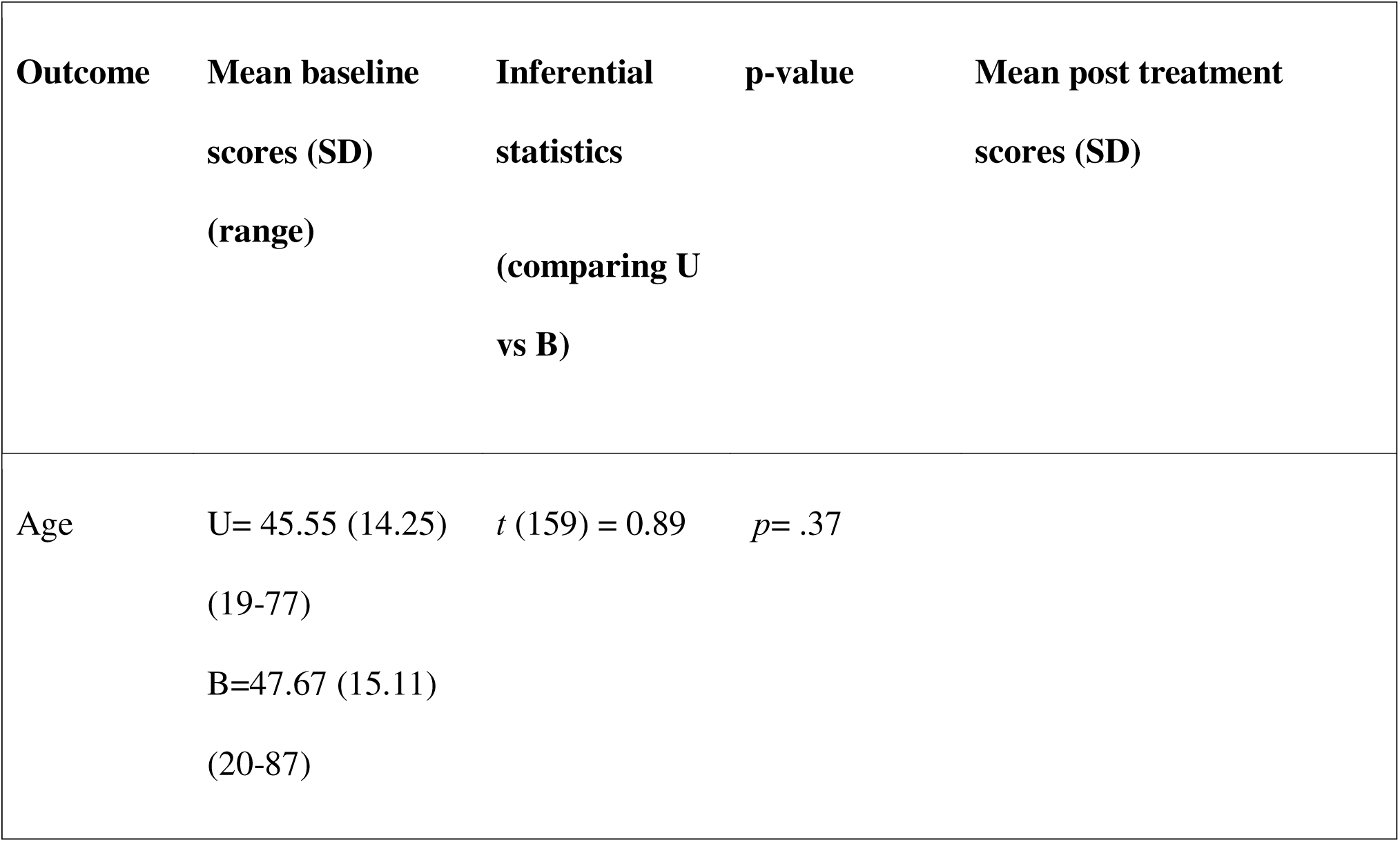
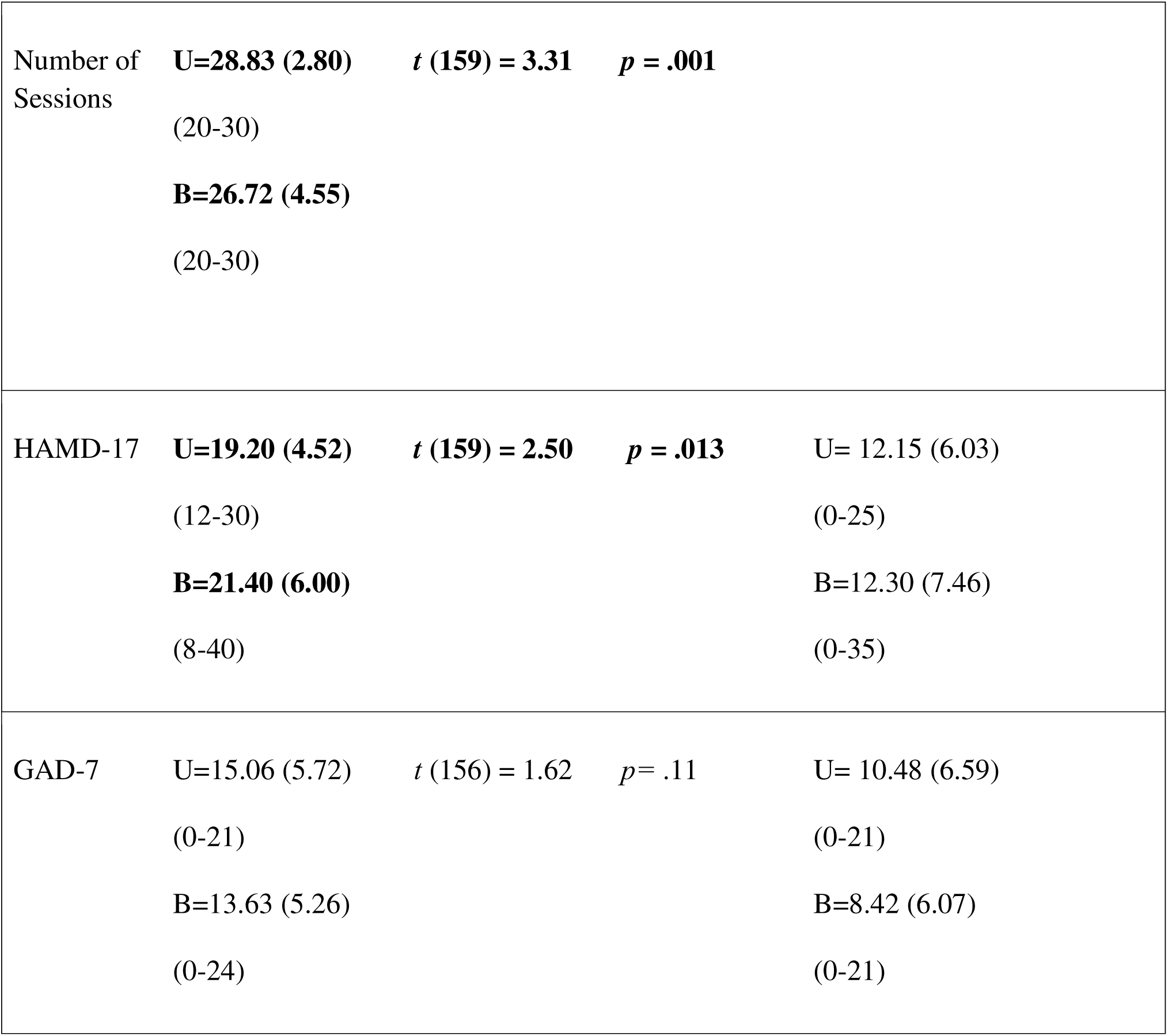
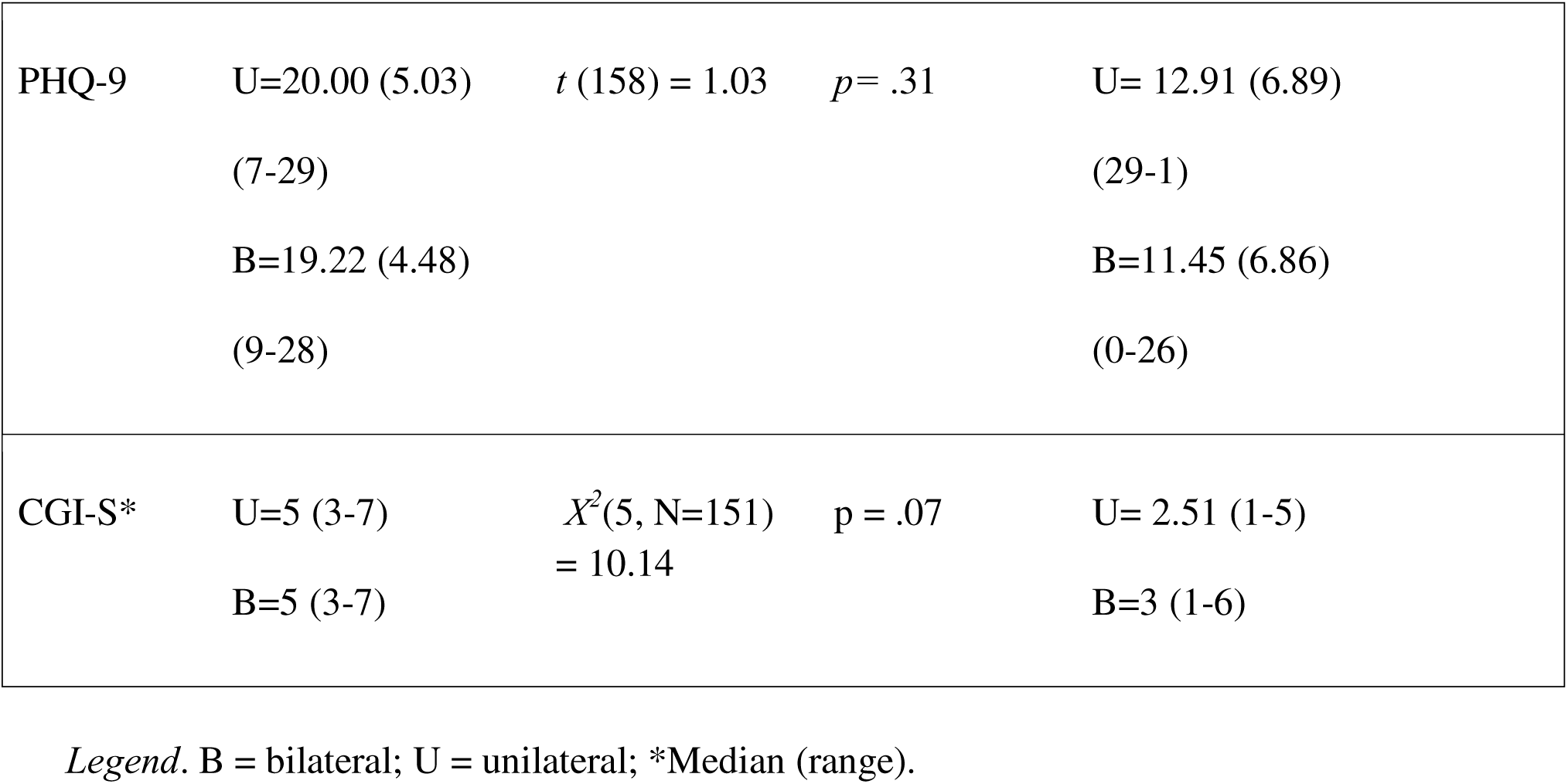
Unilateral (liTBS) and Bilateral (bsTBS) Patient Comparison on Demographics and Baseline and Post Treatment Scores.

We conducted analyses to determine if there was a difference between unilateral and bilateral stimulation in the primary outcome (HAMD-17). Chi-square tests of independence revealed no significant differences between the groups in remission rates, X^2^ (1, *N* =157) = 1.15, *p*= .28, response rates, X^2^ (1, *N* =157) = 1.78, *p* = .18, and gender ratio, *X*^2^ (1, *N*=161) = .15, *p* = .70. T-tests revealed significant differences between the groups in pretreatment HAMD-17 scores, *t* (159) =2.50, *p* =.013, and number of sessions, *t* (159) = 3.31, *p* = .001. Consequently, when we accounted for these baseline differences between unilateral and bilateral treatment, an ANCOVA revealed no differences in post-treatment HAMD-17 scores, *F* (1, 153) =1.21, *p*= .27. Additionally, we examined differences between the two groups on the PHQ-9, GAD-7 and CGI-I and CGI-S while controlling for differences in the number of sessions. There were no differences found between the two groups on the PHQ-9, *F* (1, 150) = 1.55, *p=* .22, but there was a significant difference between the two groups for the GAD-7, *F* (1,148) = 3.95, *p* =0.049. Lastly, there was no difference between the groups on CGI-I, *X*^2^ (1, N=148) = 11.95, *p= .06.* Results of the MANOVA examining side effects yielded statistically significant differences between unilateral and bilateral stimulation for post headache and pain or discomfort, Wilks’ Λ= .625, *F* (11, 146) =7.97, *p*=< .001, partial η2=.38. Individual ANOVA tests were conducted for side effects of post headache and pain or discomfort, finding significant differences between the two groups, with unilateral patients experiencing more post headache and pain or discomfort relative to bilateral patients, *F* (1, 156) =68.11, *p* <.001 and *F* (1, 156) = 31.90, *p* < .001, respectively. See Table 2 .

**Table 2.** Differences in Side Effects Between Unilateral and Bilateral Stimulation.

| **Side effect** | **Inferential statistics** | **p-value** | **Side effect experienced at least once during treatment course (U)** | **Side effect experienced at least once during treatment course (B)** |
| --- | --- | --- | --- | --- |
| **Post headache** | $F = (1, 156) = 68.11$ | $p < .001$ | 85.94 % | 64.89% |
| **Muscle contractions** | $F = (1, 156) = 3.57$ | $p = .06$ | 100% | 100% |
| **Pain or discomfort** | $F = (1, 156) = 31.90$ | $p < .001$ | 95.31 % | 85.11% |
| **Scalp**<br>**irritation** | $F = (1, 156) = .41$ | $p = .52$ | 46.88 % | 46.81% |
| **Syncope** | $F = (1, 156) = 3.42$ | $p = .07$ | 0% | 3.19% |
| **Heart rate**<br>**drop over 15**<br>**bpm** | $F = (1, 156) = .03$ | $p = .87$ | 39.06% | 36.46% |
| **Heart rate**<br>**rise over 15**<br>**bpm** | $F = (1, 156) = 1.53$ | $p = .22$ | 9.38% | 13.54% |
| **Post diastolic**<br>**drop over 10**<br>**mm hg** | $F = (1, 156) = .96$ | $p = .33$ | 71.88% | 73.96% |
| **Post diastolic**<br>**rise over 10**<br>**mm hg** | $F = (1, 156) = .88$ | $p = .35$ | 59.38% | 54.17% |
| **Post systolic**<br>**drop over 10**<br>**mm hg** | $F = (1, 156) = 3.32$ | $p = .08$ | 93.75% | 67.71% |
| **Post systolic**<br>**rise over 10**<br>**mm hg** | $F = (1, 156) = .48$ | $p = .49$ | 65.63% | 47.92% |

*Notes.* B = bilateral, U = unilateral.

Structural equation modeling revealed the expected significant associations of pre-treatment scores on post-treatment scores, but there were no main effects or moderations observed (i.e., no significant predictors of treatment outcomes). Nevertheless, the resulting model was a good fit to the data (χ2(42) = 53.85, p > .05; CFI = .98; RMSEA = .04, 90%CI [.00, .08]; SRMR = .07) (See Figure 1). Additional exploratory analyses were run using the number of comorbidities and dummy codes for depressive disorder type (major depressive disorder, persistent depressive disorder, bipolar depression), personality disorder comorbidity (present, absent) and anxiety disorder comorbidity (present, absent) as (1) predictors of post-treatment HAMD-17 scores and (2) to examine for potential interactions with pre-treatment HAMD-17 scores. There were no significant main effects or interactions and none of the model associations significantly changed with the aforementioned predictors included (p > .05) .

**Figure 1.**
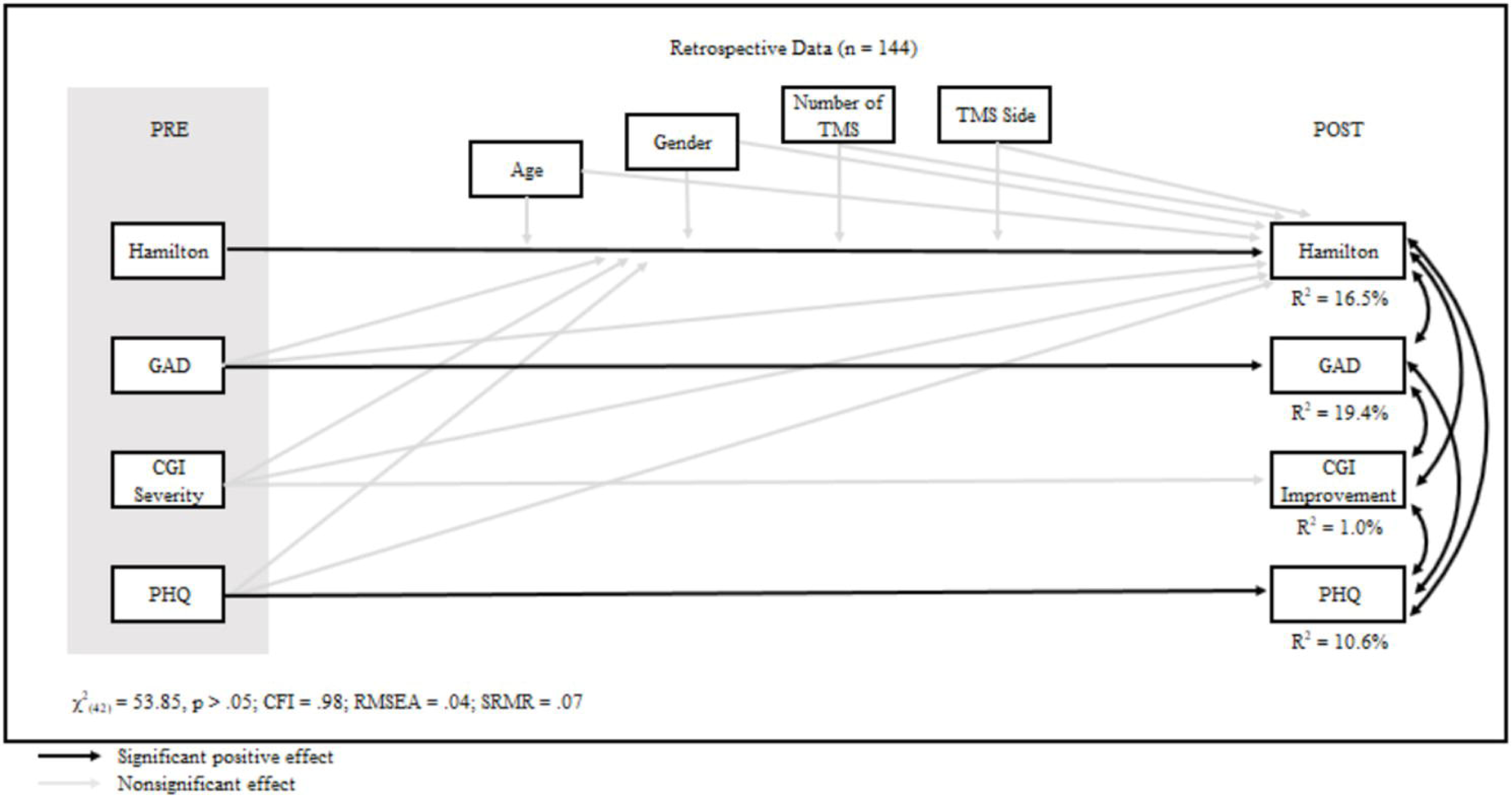
Structural equation model examining impact of predictors on treatment outcomes. Legend: CGI Clinical Global Impression Scale-Severity and Improvement; GAD Generalized Anxiety Disorder scale; Hamilton: Hamilton Depression Rating Scale-7; PHQ: Patient Health Questionnaire-9; Number of TMS: number of bsTBS or liTBS sessions; TMS side: laterality of TMS (bsTBS vs. liTBS).

Lastly, six months post TBS treatment completion, follow up data analysis was conducted on the 43% of patients who achieved response criteria: 69% of these patients did not need an additional treatment or change in medication, whereas 31% needed an additional treatment or change in medication (57.14% completed a second course of TBS treatment, 9.52% completed a course of ECT treatment, and 38.1% required a change in their medication).

## Discussion

In this study, we present observations of the effects of bsTBS, a novel version of rTMS, in a naturalistic clinical sample. First, we compared the bsTBS remission and response rates in our larger cohort of patients to those reported by Burhan et al. (2020).^6^ The analysis showed no significant differences, supporting that bsTBS remains an effective treatment option for TRD. We then compared the effects of unilateral and bilateral stimulation, accounting for baseline demographics, pre-treatment scores, and number of TBS sessions. The bilateral cohort had significantly higher pre-treatment HAMD-17 scores and a shorter treatment duration (two days less) than the unilateral group. Despite these differences, there were no significant differences in remission and response rates between the two groups. The HAMD-17 changes were also similar between the groups.

Interestingly, patients undergoing bilateral stimulation showed greater improvement on the GAD-7 scores, suggesting a potential advantage for anxiety symptoms in this group. Side effect comparisons surprisingly found that the unilateral group reported higher levels of post-treatment headache and pain or discomfort. When further examined using SEM, our analysis revealed no significant predictors of treatment efficacy. Finally, at the six-month follow-up, we found that over two-thirds of responding patients did not require additional treatment or medication change, indicating the sustained effectiveness of TBS as a treatment for TRD. Overall, our findings provide further support for the effectiveness of bsTBS in treating TRD and highlight potential differences in outcomes between unilateral and bilateral stimulation that warrant further investigation.

Our similar outcome observations to Burhan et al. (2020)^6^ are also supported by the Four-D study, which tested the non-inferiority of bsTBS to bilateral rTMS in a larger sample of geriatric patients.^5^ Relative to the Four-D study, we observed a higher depressive symptom response rate of 49%, which appears consistent with a recent meta-analysis reporting an response rate of 44.6% .^5,16^ Our results provide further support that bsTBS is safe and potentially safe in the naturalistic settings, in ages as low as 20 years.

The liTBS association with increased rate of headaches is challenging to reconcile with the greater number of magnetic pulses delivered to scalp musculature with bsTBS. Despite this difference, our observed headache rates appear higher than those in a recent meta-analysis of studies including conventional rTMS trials^17^, and similar to rates reported when comparing bsTBS to bilateral rTMS.^5^ These findings suggest that rates of TBS-induced headaches in the naturalistic setting are likely similar to those in controlled settings. Though at least one case of liTBS-induced seizure has been reported, favourable seizure safety data exists on more intensive rTMS approaches.^18–20^ The absence of seizures in our sample additionally supports the rarity of this severe side effect under the bsTBS configuration and extends the available side effect data of bsTBS to include younger age groups. That no reliable pre-treatment predictors were identified for depressive symptom response suggests that larger studies and/or updated statistical or machine learning methods would be needed to identify depressive symptom response predictors. Additionally, future studies could also examine for predictors of remission or HDRS-17<3, which may present a more binary test of depressive episode resolution than treatment response.^21^

In considering the association of bsTBS with decreased anxiety symptom ratings, we note a relatively recent meta-analysis of rTMS for anxiety, which included several studies utilizing right sided stimulation.^22^ Results from a previous post-traumatic stress disorder (PTSD) naturalistic study from our laboratory suggests bsTBS is associated with an anxiolytic effect size of close to 1.5 standardized units of anxious symptom rating improvement.^7^ These effects appear somewhat lower than the aforementioned rTMS-anxiety meta-analysis results (close to 2 units), and higher than a recent liTBS naturalistic study showing an anxiolytic effect size of 0.9.^22–23^ Notably, the same meta-analysis from Cirillo et al. (2019) showed that both inhibitory and excitatory stimuli can be associated with anxious symptom reductions. These studies suggest that adding rcTBS to liTBS (i.e. bsTBS) could provide a potential anxiolytic effect beyond liTBS alone. Introducing additional rTMS methods to treat comorbid anxiety may benefit patients with TRD, as anxiolytic effects may be associated with suicidality reduction and depressive symptom remission. ^24–25^ However, it is also important to note that bsTBS was not found to be effective for mixed mood episodes.^26^ These results suggests that clinicians need to carefully assess the phenomenological overlap between anxious depression and irritable mixed mood episodes when selecting the appropriate TRD treatment.^27^

Our 6-month post-treatment data appear similar to sequential treatment outcome data in the STAR*D trial, where close to a third to one half would not respond to a given treatment. ^28^ Our post-treatment results serve to partially extend the findings of recent multicentre studies, by highlighting the extent of long-term TBS outcomes in the naturalistic setting.^11^

While bsTBS is designed to reduce frontal asymmetry, the bsTBS mechanism requires further characterization.^29^ Notably, analyses of the impact of bsTBS on the cognitive control network^30^ and on executive functioning^31^ appeared to yield negative results. In contrast, other studies suggested that bsTBS significantly modulates the salience network^32^ and that the distribution of 5HT1A receptors could partially explain the variance of bsTBS outcomes.^33^ Nonetheless, some of those studies, while negative, provide additional support for the safety of bilateral stimulation in an array of brain regions. It is also important to note the growing importance of utilizing neuronavigation in mechanistic studies^34^, especially given the sensitivity of inhibitory vs. excitatory effects to between-location and within-location effects^35–39^, relative to stimulus parameters. ^40^ This subset of a growing list of neuronavigation studies suggests a greater need for spatial TBS precision to improve the comparative studies between stimulus site and effect profile.

We note several limitations of this study. The naturalistic study is retrospective and necessarily open-label, and non-randomized, so the results cannot demonstrate how our centre’s TRD outcomes changed as a result of transitioning from bsTBS to liTBS. Similarly, a placebo effect could lead to artificially enhanced antidepressant and anxiolytic observations. When considering the potential for the unblinded study to introduce bias in rTMS studies, one may consider recent research on potential sources of the placebo effect ^41–42^ and the projected magnitude of the placebo effect in antidepressant treatments.^43–44^ We also acknowledge the potential differences in TBS parameters from other studies, and we refer readers to our earlier study for a larger discussion of parameter differences.^6^ Another noteworthy limitation within the scope of our naturalistic approach is the single-centre design. Notably, statistics on repeat neuromodulation treatments may underestimate the population rate, as our data would not include neuromodulation treatment undertaken at treatment sites outside our centre. Lastly, we note that the SEM models can suggest without confirming the direction of any statistically significant association. Regardless, our work, among many retrospective, naturalistic studies, can be helpful to TRD clinics with access to TMS and varied patient populations but lacking neuronavigation technology, and suggests bsTBS as a safe and potentially effective option for TRD in a potentially representative sample of TRD patients.

## Conclusion

In conclusion, our work potentially adds further support to studies suggesting the effectiveness of liTBS and bsTBS to safely reduce depressive and anxious symptoms in the naturalistic setting.^11,6–7^ Our study extends recent findings from Bouaziz et al. (2023)^11^ by presenting outcomes from both unilateral and bilateral TBS paradigms, offering multiple outcome measures, and utilizing SEM to account for statistical artifacts such as regression to the mean. The findings also introduce observations of longer-term TBS outcomes in the naturalistic setting. Further retrospective analysis suggested an increased anxiolytic effect for bsTBS, which could reflect the inclusion of right hemisphere stimulation. These results highlight that bsTBS can remain a feasible option for patients and providers to consider in the lengthy and sequential process of care for TRD, perhaps especially for TRD patients with significant comorbid anxious symptoms.

## Data Access

To protect participant privacy, the data analyzed during the current study are available from the corresponding author on request.

## Acknowledgments

We would like to thank Elizabeth Russell, Mental Health Librarian at Parkwood Institute.

